# Prognostic Imaging Biomarkers for Diabetic Kidney Disease (iBEAt): Study protocol

**DOI:** 10.1101/2020.01.13.20017228

**Authors:** Kim M Gooding, Chrysta Lienczewski, Massimo Papale, Niina Koivuviita, Marlena Maziarz, Anna-Maria Dutius Andersson, Kanishka Sharma, Paola Pontrelli, Alberto Garcia Hernandez, Julie Bailey, Kay Tobin, Virva Saunavaara, Anna Zetterqvist, David Shelley, Irvin Teh, Claire Ball, Sapna Puppala, Mark Ibberson, Anil Karihaloo, Kaj Metsärinne, Rosamonde Banks, Peter S Gilmour, Michael Mansfield, Mark Gilchrist, Dick de Zeeuw, Hiddo J.L. Heerspink, Pirjo Nuutila, Matthias Kretzler, Matthew Wellberry-Smith, Loreto Gesualdo, Dennis Andress, Nicolas Grenier, Angela C Shore, Maria F. Gomez, Steven Sourbron, iBEAt investigators

## Abstract

Diabetic kidney disease (DKD) is traditionally classified based on albuminuria and reduced kidney function (estimated glomerular filtration rate (eGFR)), but these have limitations as prognostic biomarkers due to the heterogeneity of DKD. Novel prognostic markers are needed to improve stratification of patients based on risk of disease progression.

The iBEAT study, part of the BEAt-DKD consortium, aims to determine whether renal imaging biomarkers (magnetic resonance imaging (MRI) and ultrasound (US)) provide insight into the pathogenesis and heterogeneity of DKD (primary aim), and whether they have potential as prognostic biomarkers in DKD progression (secondary aim).

iBEAT is a prospective multi-centre observational cohort study recruiting 500 patients with type 2 diabetes (T2D) and eGFR > 30ml/min/1.73m^2^. At baseline each participant will undergo quantitative renal MRI and US imaging with central processing for MRI images. Blood sampling, urine collection and clinical examinations will be performed and medical history obtained at baseline, and these assessments will be repeated annually for 3 years. Biological samples will be stored in a central laboratory for later biomarker and validation studies. All data will be stored in a central data depository. Data analysis will explore the potential associations between imaging biomarkers and renal function, and whether the imaging biomarkers may improve the prediction of DKD progression rates.

Embedded within iBEAT are ancillary substudies that will (1) validate imaging biomarkers against renal histopathology; (2) validate MRI based renal blood flow against water-labelled positron-emission tomography (PET); (3) develop machine-learning methods for automated processing of renal MRI images; (4) examine longitudinal changes in imaging biomarkers; (5) examine whether the glycocalyx, microvascular function and structure are associated with imaging biomarkers and eGFR decline; (6) a pilot study to examine whether the findings in T2D can be extrapolated to type 1 diabetes.

The iBEAT study, the largest DKD imaging study to date, will provide invaluable insights into the progression and heterogeneity of DKD, and aims to contribute to a more personalized approach to the management of DKD in patients with type 2 diabetes.

## INTRODUCTION

### The BEAt-DKD project

Diabetic kidney disease (DKD) is the leading cause of end stage renal disease (1,2). It is currently estimated that approximately 20-40% of people with diabetes will develop DKD (3), and this is expected to rise in the future. With the global increase in the prevalence of diabetes (4), particularly type 2 diabetes, DKD is reaching epidemic proportions, with health and quality of life implications (e.g. increased risk of cardiovascular mortality) for the individual (5). Even with current approaches to management of diabetes and renin-angiotensin-aldosterone system blockade, there is still large residual risk in DKD (6).

DKD is routinely classified clinically based on albuminuria and reduced kidney function (estimated glomerular filtration rate (eGFR)). Albuminuria is traditionally viewed as a hallmark of diabetes related kidney damage. However, there are limitations of using albuminuria to classify DKD, which include the need for multiple measurements to mitigate spurious results due to factors such as infection and physical activity. Additionally, the heterogeneity of DKD is increasingly recognised, as reflected, for example, by the disparity in DKD progression (fast versus slow DKD progression) and by patients with declining kidney function but normoalbuminuria. For example, 51% of participants in the UK Prospective Diabetes Study whose eGFR declined below 60 ml/min/1.73 m^2^ had normoalbuminuria (7). This heterogeneity in DKD highlights the need for novel biomarkers and a more personalized medicine-based approach to managing DKD.

The fundamental aim of the Biomarker Enterprise to Attack DKD (BEAt-DKD) consortium is to increase our understanding of the pathogenesis and heterogeneity of DKD, enabling the identification of novel biomarkers and treatment targets, to facilitate a more personalized medicine-based approach to managing DKD and increase the efficiency of clinical trials (8).

### Imaging biomarkers for DKD

Cross-sectional imaging, in particular MRI and US, is increasingly proposed as an alternative source of biomarkers to inform chronic kidney disease (CKD) management (9,10). An important example is the qualification by the Food and Drug Administration (FDA) and the European Medicines Agency (EMA) of Total Kidney Volume (TKV) as a prognostic enrichment biomarker for Autosomal Dominant Polycystic Kidney Disease (ADPKD) – one of only a handful of clinical biomarkers approved by the FDA so far (11,12). In recent years the interest is increasingly moving towards advanced MRI and US techniques that are sensitive to structural and functional tissue characteristics such as perfusion, oxygenation, blood flow, glomerular filtration, tubular flow, fibrosis, inflammation, metabolism and tissue composition. Additional utility derives from the fact that these characteristics can be measured separately for left and right kidney, for cortex and medulla, or to map functional and structural heterogeneity within those areas.

A number of preclinical and single-centre clinical studies have indicated a potential utility of MRI and US biomarkers in DKD specifically. For instance, US-based measurements of kidney volume have suggested that kidney enlargement is associated with poorer outcomes in early and advanced DKD, despite the often better GFR of larger kidneys (13–15). A possible explanation is that hypertrophy indicates a sustained state of primary or secondary hyperfiltration and associated damage due to intraglomerular pressures. A mechanistic study suggested that the MRI method BOLD (Blood Oxygenation Level Dependent MRI) can highlight areas at risk of ischemic damage due to oxygen depletion after sustained hyperfiltration (16), and a recent clinical study has confirmed that the BOLD signal is predictive of CKD progression (17). Some MRI biomarkers derived from diffusion-weighted MRI are sensitive to renal fibrosis (18,19) and can identify microstructural changes after sustained hyperfiltration (20), though the clinical potential of MRI measures of fibrosis and microstructure remains to be confirmed (21). Kidney perfusion and glomerular filtration can be measured with MRI. Renal blood flow has been shown to correlate with eGFR in DKD (22). Other n-renal imaging biomarkers characterising general risk factors for diabetes and its associated complications may be relevant in this context as well and can easily be measured in the same MRI scan session, such as liver and pancreatic fat fraction (23).

### iBEAt study aims and objectives

The aim of iBEAt is to evaluate the evidence for the utility of imaging biomarkers in DKD in a large cohort of heterogeneous type 2 diabetes patients, in the early stages of DKD where there is high potential for effective interventions to slow the rate of DKD progression.

The key hypotheses are that (1) imaging-based biomarkers of DKD provide additional information on the pathogenesis and histological and clinical heterogeneity of DKD compared to biomarkers sourced from samples or physical exams, and (2) that changes in imaging biomarkers precede increases in albuminuria and decline in kidney function as measured by eGFR. As a result, we expect imaging biomarkers to improve the identification of DKD patients at risk of rapid decline in kidney function, either when used alone or combined with clinical data / biological fluid biomarkers.

An additional aim of the iBEAT study is to establish a biobank of biological samples (blood- and urine-based) from well-characterised patients for use within the BEAt-DKD programme and future DKD studies. This will facilitate biomarker discovery studies using novel blood- and urine-based biomarkers and may serve as the foundation for a comprehensive multi-scale phenotyping strategy linking data from blood, urine, tissue, microvascular assessments, imaging, physical measurements and medical histories.

The specific study objectives are:

- Primary objective: To examine whether renal imaging biomarkers are associated with severity of DKD as defined using classical biomarkers of DKD, albuminuria and eGFR, in individuals with type 2 diabetes and eGFR > 30 ml/min/1.73m^2^.
- Secondary objective: To examine whether renal imaging biomarkers are associated with changes in renal function over time as measured by eGFR over a 3-year period.

### Overview of iBEAt study design and organisation

iBEAT (registered at clinicaltrials.gov under NCT03716401) is a prospective observational study that will enrol 500 participants, stratified into six subgroups based on three albumin-to-creatinine ratio (ACR) and two eGFR categories, with type 2 diabetes (T2D) and eGFR greater than 30 mL/min/1.73m^2^ across five European centers.

A schematic overview of the study assessments is presented in Table 1. At baseline, each participant will undergo comprehensive renal imaging (MRI and US), biological sample collection (blood and urine), physical measurements and their medical history will be collected. They will then be invited back annually for 3 years, where all measurements except the imaging will be repeated. Participants can then be followed remotely, through medical notes and/or questionnaires, for a further 15 years.

**Table 1.** Overview of the study showing type of data collected (rows) for each time point (columns).

| Protocol Details | Screening | Baseline | Y1 | Y2 | Y3 |
| --- | --- | --- | --- | --- | --- |
| Time Window | Day 0 | 0-3m | 1y±3m | 2y±3m | 3y±3m |
| Informed Consent | X |  |  |  |  |
| Demographics | X |  |  |  |  |
| Clinical Information |  | X | X | X | X |
| Local lab value collection | X | X | X | X | X |
| Blood Collection |  | X | X | X | X |
| Urine Collection (random) | X |  |  |  |  |
| Urine Collection (1st morning & additional void) |  | X | X | X | X |
| MRI |  | X |  |  |  |
| US |  | X |  |  |  |
| DNA & RNA Sample |  | X | X | X | X |

The organisation of iBEAT is shown in Figure 1. The study is led by the coordinating centre in Leeds with a co-lead in Exeter and a study manager in Michigan, currently there are 5 recruiting centers (University of Leeds, University of Exeter Medical School, University of Bari, University of Bordeaux and University of Turku), a central laboratory (Lund University) and a central data repository (Swiss Institute of Bioinformatics (SIB)). All ethical and relevant local approvals are in place at each recruiting site. As a BEAt-DKD work package the study is supported by the BEAT-DKD consortium Steering Committee and an external Scientific advisor.

**Figure 1.**
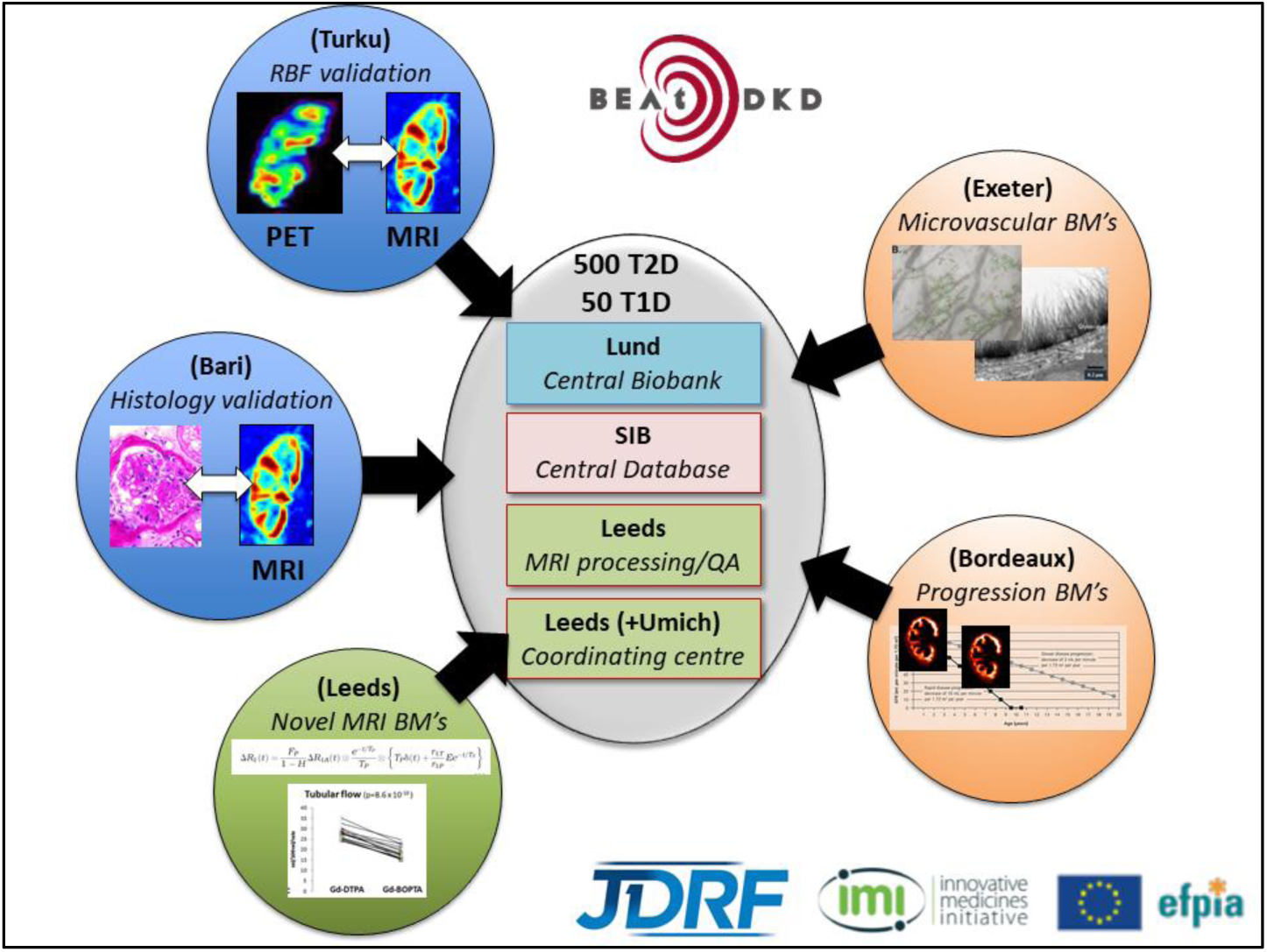
iBEAt study organigram showing central roles (grey ellipse) and recruiting sites with ancillary studies (circles). BMs = biomarkers; QA = quality assurance; RBF = renal blood flow; PET = Positron-emission tomography; SIB = Swiss Institute of Bioinformatics; Umich = University of Michigan.

## METHODS

### iBEAt participants

The iBEAt study will recruit participants with a diagnosis of type 2 diabetes, eGFR greater than 30 mL/min/1.73m^2^, aged between 18-80 years, who are able to give informed consent, and do not satisfy any of the exclusion criteria. The exclusion criteria are listed in Table 2 (see also supplement 3.0).

**Table 2.**
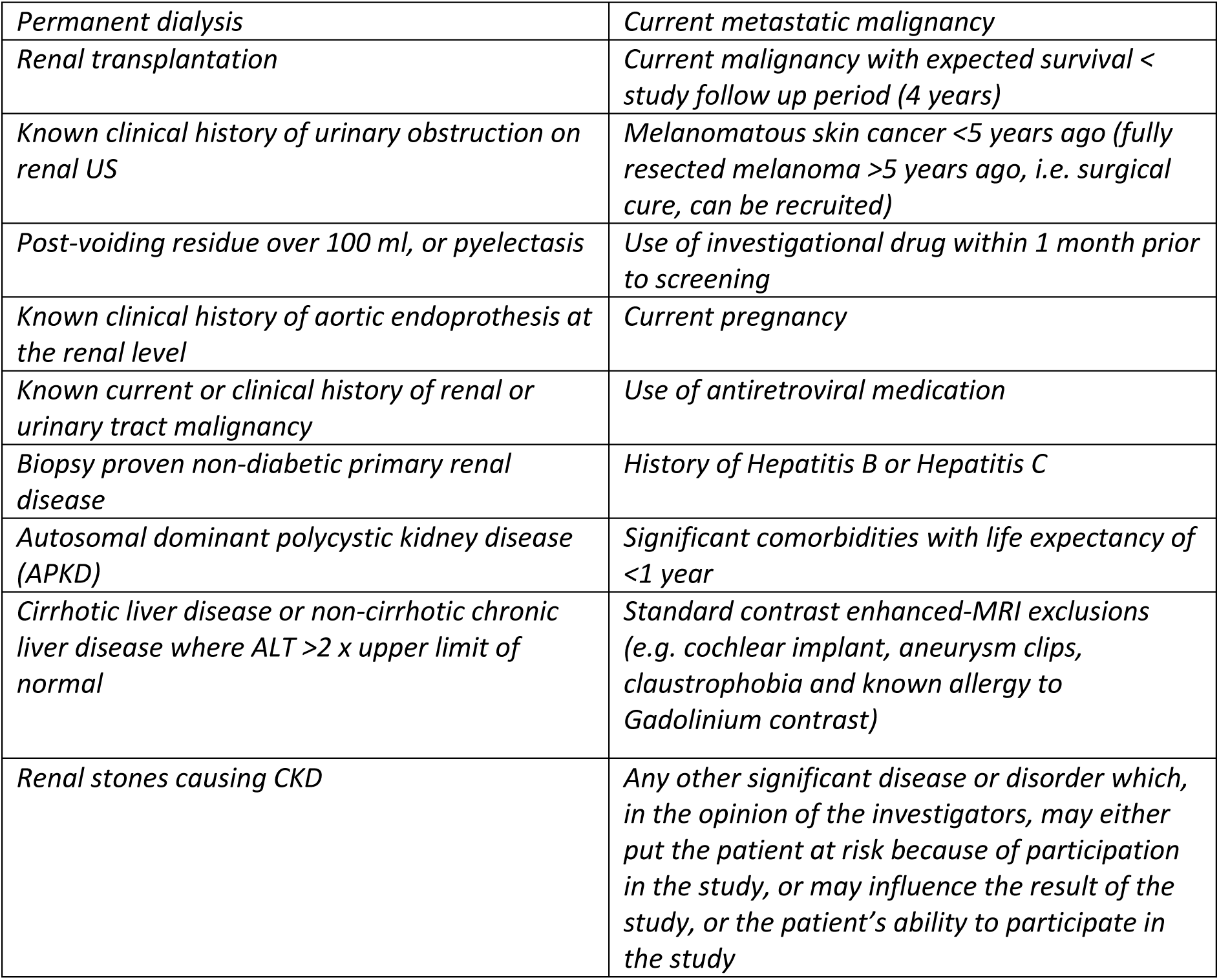
List of iBEAt exclusion criteria (see also supplement 3.0).

|  |  |
| --- | --- |
| *Permanent dialysis* | *Current metastatic malignancy* |
| *Renal transplantation* | *Current malignancy with expected survival < study follow up period (4 years)* |
| *Known clinical history of urinary obstruction on renal US* | *Melanomatous skin cancer <5 years ago (fully resected melanoma >5 years ago, i.e. surgical cure, can be recruited)* |
| *Post-voiding residue over 100 ml, or pyelectasis* | *Use of investigational drug within 1 month prior to screening* |
| *Known clinical history of aortic endoprosthesis at the renal level* | *Current pregnancy* |
| *Known current or clinical history of renal or urinary tract malignancy* | *Use of antiretroviral medication* |
| *Biopsy proven non-diabetic primary renal disease* | *History of Hepatitis B or Hepatitis C* |
| *Autosomal dominant polycystic kidney disease (APKD)* | *Significant comorbidities with life expectancy of <1 year* |
| *Cirrhotic liver disease or non-cirrhotic chronic liver disease where ALT >2 x upper limit of normal* | *Standard contrast enhanced-MRI exclusions (e.g. cochlear implant, aneurysm clips, claustrophobia and known allergy to Gadolinium contrast)* |
| *Renal stones causing CKD* | *Any other significant disease or disorder which, in the opinion of the investigators, may either put the patient at risk because of participation in the study, or may influence the result of the study, or the patient's ability to participate in the study* |

iBEAt will recruit across six strata defined by the A1-A3 albuminuria range (normo-, micro- and macroalbuminuria) and the (G1+G2)-G3 eGFR range. In line with the National Kidney Foundation guidelines (24), albuminuria will be classified using two (or three) independent values of ACRs measured within a 3-month period: one at the screening visit, one at the baseline study visit, and a third if the classification differs between the first two samples. We define normo-, micro- and macro-albuminuria as an ACR of <2.5, 2.5–25, >25 mg/mmol for men, respectively, and as ACR <3.5, 3.5–35, >35 mg/mmol for women, respectively.

### Study enrolment

Following the provision of written informed consent at the start of the screening visit, potential participants undergo a number of health assessments including, for example, medical history, random spot urine collection for assessment of ACR and a blood sample for the assessment of eGFR (unless these have been performed in the last 3 months) to assess eligibility for the study.

### Baseline study assessments

#### Participant preparation

Fasting blood samples are taken in the morning following an overnight fast. Medications may be withheld or altered on the day of study visit to ensure participant wellbeing (e.g. omitting morning insulin injection to maintain blood glucose levels) and integrity of the study. A point of care glucose measurement will be performed upon participant arrival and the visit will be cancelled if glucose levels are below 3.5mmol/L or if the participant reports a symptomatic hypoglycaemic event on the morning of the visit. All other assessments are performed following a standardised meal. Participants are required to be on stable diabetes and hypertension related treatment (though dose changes to current medications are allowed) for the 3 months prior to study assessments. The study visit will include a checklist to record the adherence to instructions (supplement 3.1).

#### MRI biomarkers

59 primary MRI biomarkers will be recorded (see supplement 1.0 for a full list), characterising general body composition (e.g. visceral fat volume, pancreatic and liver fat fraction), renal morphology (e.g. parenchymal volume, cortical thickness), renal tissue structure (e.g. MR relaxation times, apparent water diffusion coefficient), renal hemodynamics (e.g. cortical perfusion, renal artery blood flow), filtration (e.g. single-kidney GFR, filtration fraction). All MR scanning is performed at 3T on Siemens, Philips and General Electric scanners. MRI data are uploaded on a central XNAT database hosted by SIB and quality controlled within 48hrs by the central processing site in Leeds.

The MRI protocol takes approximately 1hr and 10mins and involves the injection of a quarter dose of clinical macrocyclic MRI contrast agent. The protocol was first developed on the reference Siemens scanner in Leeds using an iterative optimisation guided by the NIST (National Institute of Standards and Technology) phantom and healthy volunteers. The resulting final protocol was then characterised on each MRI vendor using a repeatability study in healthy volunteers to determine within-site variability (5 volunteers with 4 scans each). The NIST phantom is scanned at regular intervals in all sites to check for between-site calibration. Full details of the MRI acquisition protocol on the 3T Siemens reference scanner in Leeds can be found in supplement 1.1.

#### Renal ultrasound

Kidney size will be non-invasively determined from longitudinal and transversal images of each kidney. Resistive index (RI), indicator of the resistance to flow within the kidney, will be determined from three measurements in each kidney (upper, mid and lower poles). The mean of the three measurements will represent RI for each respective kidney. A list of US biomarkers are provided in the supplement 3.7 and the Standard Operating Procedures (SOPs) for US scanning are in supplement 1.2.

#### Blood and urine sampling

Fasting blood samples (∼70mls designated for iBEAT central requirement) will be collected from each participant for participant characterisation and biomarker analysis. Glycated haemoglobin (HbA1c), full blood count and fasting glucose assessments will be performed locally (supplement 3.4). The remaining plasma and serum samples will be processed and stored following a standardised protocol (see supplemental 2.1-2.3). A first morning urine void and one additional morning void (same day) are collected by all participants. A small proportion of the first morning void is sent to the local laboratories for ACR assessment. The remainder of the first morning and second void are then processed and stored following a standardised protocol (see supplemental 2.1-2.3).

The standardised sample collection and processing protocol, informed by Provalid and Neptune trials (25,26), was developed to maximise the utility of stored samples for future biomarker analysis (e.g. lipidomics, RNA analysis, urinary vesicles and urinary sediment) within BEAt-DKD and to form a biobank for future DKD related studies. A separate check is performed to confirm that all samples are collected and processed according to protocol (supplement 3.8).

The central biochemical laboratory is located at the Clinical Research Centre (CRC) facility in Malmö (University of Lund). The central lab will prepare and distribute kits with sample collection and processing materials for each patient labelled and barcoded with the study ID. Each kit comprises of 66 storage tubes per patient. The samples will be temporarily stored at each recruiting site, with regular shipments returning them to the biobank in Malmö. Samples will be stored under secure conditions and monitored with a dedicated electronic sample tracking system (Laboratory Information Management Systems). A small volume of blood and urine will be analysed in Malmö for known clinical biomarkers (e.g. renal function (serum creatinine, cystatin C, potassium and albumin), lipid profile (total cholesterol and sub-fractions, triglycerides) and c-reactive protein) at the central laboratory, according to standardised methods, and the remainder will be stored for future analyses by BEAt-DKD investigators. The samples will also remain available for secondary research provided approval is granted by the iBEAT steering committee.

#### Physical examination

The core physical examination assessments include blood pressures (sitting and standing blood pressures) and anthropometrics (height, weight, waist and hip circumference) assessments. See supplement 3.2 for details.

#### Medical history

A detailed medical history (including, for example, current medications, smoking history and presence of co-morbidities) is also collected. See supplements 3.3 and 3.6 for data fields that are captured.

#### Routine lab data

Routine local laboratory data will be captured to aid in the interpretation of the results by tracking temporal changes at a finer time scale than the yearly follow-ups. Only laboratory values available for clinical indication will be captured at this time. Supplement 3.4 lists the data fields to be captured but missing data from the local chart is not deemed a protocol violation.

### Follow-up study assessments

The biological sampling protocol, medical history and physical examination will be repeated at 1, 2 and 3 years (± 3months) following study enrolment. For participants who are unable to attend the local research centre for an annual follow-up visit but are still willing to participate in the study an update on their medical history will be collected via direct communication with the participant and / or by accessing their available medical records.

### Data management

Clinical images, associated data and metadata will be stored using the XNAT platform (www.xnat.org) hosted on the dedicated BEAt-DKD server at SIB. Clinical study data will be managed using REDCap (www.project-redcap.org), also installed on the dedicated BEAt-DKD server. All variables will be recorded on iBEAt central Clinical Record Folders (CRF’s – see supplements 3.0-3.8) and uploaded onto the central RedCap instance. It is envisaged that the iBEAT clinical study will be set up as a federated node enabling remote analysis of the data generated in the future, and integration of the iBEAt data with other datasets collected in BEAt-DKD.

### Statistical considerations

#### Sample size

The sample size calculation for a study such as this one would be complex and unlikely to be accurate. Thus, we opted for a more pragmatic approach, and arrived at the sample size by considering the scientific and feasibility aspects of the study. Specifically, in considering the sampling design, we were mainly concerned with whether it would allow us to answer the scientific questions we have and whether it would be feasible to carry out. We opted for a stratified sampling design, as we were interested in evaluating the association between imaging renal biomarkers and DKD in type 2 diabetes patients in various stages of DKD, as measured by ACR and eGFR, widely used biomarkers of DKD. This ensures that we will have a reasonable sample size for all combinations of ACR (A1, A2, A3) and eGFR (G1+G2, G3), even those that are typically rare, specifically A1 and G3, as well as A3 and G1+G2. In terms of feasibility, we considered the number of imaging facilities available to us and the estimated rate of recruitment in each for each strata of our sample. Based on these considerations we arrived at a stratified sample of 500 patients, 4 groups of 92 and two groups of 66. See Table 2 in Supplement 5.0 for details.

#### Statistical analysis plan

We will begin by describing our data, comparing the descriptive statistics of all covariates across the study centers, assessing the bivariate relationships between sets of related covariates. We will then perform a cross-sectional analysis using data collected at baseline, as well as a longitudinal analysis using the imaging data collected at baseline and blood and urine markers collected over the three years. Given the large number of covariates in our study, we will use variable selection and regularization methods such as LASSO (27) and the elastic net regularization (28). The modelling will be done appropriately for a given setting, with linear models for the cross-sectional analyses, and linear mixed effects models for the longitudinal analysis. The analyses will be performed separately in each strata and, whenever possible, accounting for the stratification in the modelling, as we expect there to be effect modification in the potential associations we will be estimating across the strata. We will adjust for multiple comparisons as needed. If we identify any renal imaging biomarkers as having promise as early markers of DKD progression, we will use them in risk prediction and evaluate their predictive accuracy, as measured by prediction error, receiver operating characteristics curve (ROC) and the area under the ROC (area under the curve, AUC), using cross-validation.

We plan to take a conservative approach in all of our analyses, meaning that we will be careful with the number of models we fit and statistical tests we perform, and will treat our analyses as exploratory and hypotheses generating, rather than hypothesis testing.

### Ancillary studies

Building on the strengths and interests across the iBEAt participating centres, six ancillary studies have been incorporated within the central iBEAt study. Participants taking part in the ancillary studies will be recruited from the central iBEAt study at the relevant sites.

#### Ancillary study 1

To examine whether MRI and US based imaging biomarkers correlate with histopathological markers of DKD and discriminate different renal lesions in this T2D cohort. For this ancillary study, led by Bari University, iBEAt participants will undergo a renal tissue core biopsy (n=100). All biopsies will be digitalized and characterized by Light Microscopy (hematoxylin-eosin, periodic acid-Schiff, silver methenamine, and Masson’s trichrome), Immunofluorescence Microscopy (with the use of antisera against IgG, IgM, IgA, C3, C4, C1q and fibrinogen) and Electron Microscopy. Glomerular and vascular lesions, interstitial cell infiltrate, fibrosis and tubular atrophy will be quantified (29). Samples will also be processed and stored for later biomarker discovery. The procedures for processing, storing and capturing meta-data regarding the renal biopsy tissue are described in more detail in supplement 4.0.

#### Ancillary study 2

To examine whether MRI-based measurements of renal blood flow correlate with H2O-positron emission tomography (PET) renal perfusion measurements, validating the MRI based measurements against the standard PET perfusion measurements. For this ancillary study, led by Turku University, a direct comparison of MRI and PET-based measurements of renal blood flow will be performed in a cohort of iBEAT participants. Renal perfusion will be assessed during hyperaemia with both systems.

#### Ancillary study 3

To develop machine-learning methods for automated or semi-automated processing of multiparametric renal MRI. In its current form the generation of biomarkers from complex functional MRI scans involves significant manual intervention as well as automated but slow iterative optimisation methods. In this study, led by Leeds University, a subset of the iBEAt data will be used as training data to develop an ideally automated approach for image processing, which will then be validated on the remaining test data against the manual results.

#### Ancillary study 4

To investigate the longitudinal changes in MRI and US based biomarkers, compare them against changes in eGFR and other known markers, and determine whether changes in imaging biomarkers precede DKD progression as assessed by eGFR decline. For this study a cohort of 100 patients will receive repeat MRI and US after 2 years, and changes in imaging biomarkers over that period will be correlated against changes in eGFR and other assessments.

#### Ancillary study 5

To examine whether the glycocalyx, microvascular function and structure (retinal and skin) are (1) altered in microalbuminuria; (2) associated with DKD progression as assessed by eGFR decline and (3) are associated with novel MRI and US imaging DKD biomarkers. For this study, led by University of Exeter, iBEAt participants will also undergo comprehensive microvascular assessments (including non-invasive estimation of sublingual endothelial glycocalyx integrity, retinal vascular oxygenation and skin maximum hyperaemia) at baseline and at 2 years follow-up.

#### Ancillary study 6

A pilot study to examine whether the findings in T2D can be extrapolated to Type 1 diabetes. In this ancillary study a cohort of 50 patients with Type 1 diabetes will be assessed using the same procedures as the Type 2 cohort and observed findings / trends will be compared across the two populations.

### Patient and public involvement

Patient and public involvement and engagement is a significant component of the iBEAt study. Potential participants have played an important role in iBEAt, reviewing the protocol to ensure the feasibility of the study design (core and ancillary studies) as well as contributing to the development of patient facing documents (e.g. patient information sheets), ensuring that they are clear and informative. Participants within the iBEAt study will play an integral role in the dissemination of the study results to the wider, non-expert population. Within the BEAt-DKD consortium discussions with patient representatives, ranging from experienced patient advocates to iBEAt participants, will help inform how research from the BEAt-DKD consortium is taken forward to implement a more precision medicine based approach in DKD into clinical practice; for example, validation and qualification of new biomarkers by regulatory agencies, optimising clinical study design and integration in the regulatory process of drug registration. Indeed, this has already commenced with an iBEAt participant, along with other patient representatives, attending the 2nd BEAt-DKD Stakeholders’ symposiums in April 2019 (30).

## DISCUSSION

Quantitative and functional imaging of the kidney has been an active topic of research in the MRI physics and radiology community for over two decades (31), but the last few years have seen an explosive growth in clinical interest. The first international meeting on functional renal MRI was held in 2015 and attendance has been increasing steadily ever since (32–34). In 2017, a pan-European network of researchers in renal MRI (www.renalmri.org) was funded for 4 years by the European Cooperation in Science and Technology (www.cost.eu). In 2018, Nephrology Dialysis Transplantation published a special issue on renal MRI with a clinical position statement supported by over 30 authors including leading European nephrologists (9). In the same year, in the US, the National Institute of Diabetes and Digestive and Kidney Diseases (NIDDK) at the National Institutes of Health (NIH) conducted a workshop on renal imaging to review the state-of-the-art and plan potential future endeavours [7]. Also in 2018, the UK Renal Imaging network (UKRIN) received a 3-year partnership grant to create a national infrastructure for quantitative renal MRI, and has advanced plans for a 10 year cohort study in 450 CKD patients (AFiRM study; principal investigator: Nick Selby, University of Nottingham).

iBEAt builds on these developments and is the first study to respond to the clinical need for systematically collected evidence at a larger scale and across institutions, with well-validated methods linking up the imaging findings with other sources of data so the added value can be identified. In that sense, iBEAt is inspired by the landmark study CRISP (Consortium for Radiologic Imaging Studies of Polycystic Kidney Disease) (35) - the first multi-centre cohort study exploring a quantitative MRI biomarker (TKV) in CKD and a foundation for the aforementioned FDA qualification of TKV. Like CRISP, iBEAt has built in a technical validation phase of the imaging biomarkers by including a repeatability study on all scanner types deployed in iBEAt, and by calibrating between-scanner differences through a travelling test object developed for this purpose by the National Institute of Standards and Technology (36). Also following the example of CRISP, iBEAt is committed to sharing the technical details of its imaging protocols and expertise in image processing and quality assurance – not only to facilitate the cost and setup of future studies but also to maximise alignment and future opportunities for pooling the data. An example is an ongoing collaboration with the DYNAMO consortium (https://www.duke-nus.edu.sg/about/achievements/awards/collaborative-grants) in setting up an imaging biomarker study aligned with iBEAt.

Collectively, the integration of the ancillary studies into iBEAT will provide valuable information on the pathogenesis of DKD and the clinical utility of these imaging biomarkers. Crucially, they will explore the association of renal based imaging biomarkers against histopathological markers and different histological lesions of DKD, validating the imaging biomarkers and substantiating their clinical utility. MRI renal based perfusion measurements will also be validated against H2O-PET renal perfusion measurements. The potential automation of the MRI image processing will streamline a labour intensive process, thereby increasing the clinical applicability of the assessments. The microvascular assessments, including the examination of glycocalyx integrity and endothelial function, will provide invaluable information on the pathogenesis and heterogeneity of DKD, and may well aid the identification of individuals with fast progressing DKD, for example, we hypothesise that individuals with type 2 diabetes with early signs of perturbations to the glycocalyx will be at an increased risk of DKD progression.

iBEAt has greatly benefitted in its setup from study documents and standard operating procedures (SOPs) provided by other investigators, in particular the PROVALID (26) and NEPTUNE (25) studies. In turn, iBEAt is committed to a “pay-it-forward” philosophy and will aim to share its study documentation and procedures widely for use by other investigators. iBEAt collaborators are also committed to maximise the opportunities for data sharing in order to increase the lifetime value of their research data as assets for human health and to do so timely, responsibly, with as few restrictions as possible, in a way consistent with the law, regulation and recognised good practice. Beyond data, iBEAt will aim to form a powerful resource for future biomarker discovery sources by collecting a rich collection of blood and urine samples in its central biobank. These will be made available for external investigators subject to formal application and approval by the iBEAt Steering Committee.

After a 2-year setup period the first study participant was recruited into iBEAt in October 2018. First results on technical validation of MRI methods on the reference scanner are expected at the end of 2019. The projected deadline for recruitment is 1 September 2020 and first results on the primary objective (cross-sectional analysis of baseline data) are expected to be made public in 2021. Completion of follow-up data is expected in 1 sept 2023 with results on the longitudinal analysis expected to be submitted for publication in 2024.

## Data Availability

iBEAt investigators are committed to maximise the opportunities for data sharing at study completion in order to increase the lifetime value of their research data as assets for human health and to do so timely, responsibly, in a way consistent with the law, regulation and recognised good practice.

## Supplementary material

Suppl 1.1. Table of MRI biomarkers

Suppl 1.2. MRI reference acquisition protocol on Siemens 3T

Suppl 1.3. US SOPs

Suppl 2.1. Blood & urine collection SOPs

Suppl 2.2. Blood & urine processing SOPs

Suppl 2.3. Blood & urine schematics

Suppl 3.0. CRF Screening

Suppl 3.1. CRF Adherence Checklist

Suppl 3.2. CRF Limited Clinical Exam

Suppl 3.3. CRF Medical and Family Hx

Suppl 3.4. CRF Local Study Labs

Suppl 3.5. CRF Routine Labs

Suppl 3.6. CRF Medications

Suppl 3.7. CRF Ultrasound

Suppl 3.8. CRF Biosamples

Suppl 4.0. Biopsy protocol

